# A dual antigen ELISA allows the assessment of SARS-CoV-2 antibody seroprevalence in a low transmission setting

**DOI:** 10.1101/2020.09.09.20191031

**Authors:** Sarah M. Hicks, Kai Pohl, Teresa Neeman, Hayley A. McNamara, Kate M. Parsons, Jin-shu He, Sidra A. Ali, Samina Nazir, Louise C. Rowntree, Thi H. O. Nguyen, Katherine Kedzierska, Denise L. Doolan, Carola G. Vinueas, Matthew C. Cook, Nicholas Coatsworth, Paul S. Myles, Florian Kurth, Leif E. Sander, Graham J. Mann, Russell L. Gruen, Amee J. George, Elizabeth E. Gardiner, Ian A. Cockburn, the SARS-CoV-2 Testing in Elective Surgery Collaborators

**Author notes:** These authors contributed equally (Elizabeth E. Gardiner, Ian A. Cockburn). These authors contributed equally (Sarah M. Hicks, Kai Pohl). Corresponding Author Contact Details Department of Immunology and Infectious Disease, John Curtin School of Medical Research,The Australian National University, Canberra, Australia.

## Abstract

Estimates of seroprevalence of SARS-CoV-2 antibodies have been hampered by inadequate assay sensitivity and specificity. Using an ELISA-based approach to that combines data about IgG responses to both the Nucleocapsid and Spike-receptor binding domain antigens, we show that near-optimal sensitivity and specificity can be achieved. We used this assay to assess the frequency of virus-specific antibodies in a cohort of elective surgery patients in Australia and estimated seroprevalence in Australia to be 0.28% (0 to 0.72%). These data confirm the low level of transmission of SARS-CoV-2 in Australia before July 2020 and validate the specificity of our assay.

## Introduction

Reported cases of severe acute respiratory syndrome coronavirus 2 (SARS-CoV-2) are likely to represent only a fraction of actual SARS-CoV-2 infections, as ∼40% of cases are mild or asymptomatic, or otherwise undiagnosed [1]. Detection of antibodies that recognize viral antigens specific for SARS-CoV-2 has become an important molecular sentinel of current or prior exposure to SARS-CoV-2 [2]. Measurement of antibody levels can provide information regarding the status of infection in an individual, as well as indicate the rate and extent of response to treatment and to recovery. Since a significant number of people either present with mild symptoms of COVID19 infection or are asymptomatic, serological measurements will have ongoing utility in gauging exposure and prevalence in the community [3]. Such studies will provide valuable information on the time course and longevity of antibody responses to SARS-CoV-2 [4]. Further, serological testing is likely to be valuable in the assessment of vaccine efficacy. However, analyses of seroprevalence, especially in low prevalence settings are hampered by assays with inadequate sensitivity and specificity [5].

Australia has reported low case numbers of COVID-19 per head of population compared to other developed Westernized countries, especially before the July/August 2020 outbreak in Melbourne, Victoria (Australian Department of Health). Efforts to control the spread of the virus have likely been helped by relative geographical isolation and an advanced healthcare system. However nucleic acid testing generally only reveals a fraction of the total numbers of infections thus the overall numbers of previous infections is unknown [3, 6]. Nonetheless it is likely that the total of previously infected individuals is low as a proportion of the population (<1%) and thus assessment of seroprevalence requires the use of highly sensitive and specific methodologies. In order to assess the seroprevalence of SARS-CoV-2 in Australia we therefore developed a dual-antigen ELISA assay which gave superior sensitivity and specificity compared to assays that rely on single antigens.

## Materials and Methods

### Samples and ethics statement

Collection of blood from individuals pre-2020 was carried out after provision of informed consent, using procedures approved by the Human Research Ethics Committees (HREC) of the Australian National University (2016/317) and ACT Health (1.16.011 and 1.15.015). Samples from SARS-CoV-2 positive individuals were collected after consent under the following protocols: Alfred Hospital HREC (280/14); James Cook University HREC (#H7886); ACT Health HREC (1.16.011): Charité Ethics Committee (EA2/066/20) [7]. Approval for the elective surgery study was given by Alfred Hospital Ethics Committee (339/20) and the Australian National University (2020/379). Whole blood was collected by venipuncture into an empty syringe (healthy donors) or a red capped serum vacutainer tube (patients), rested for 1 h then centrifuged (1000*g*, 10 min, 4°C) and the upper serum phase removed by aspiration to a new tube and immediately frozen. All samples were heated to 56°C for 1 h prior to analysis.

### ELISA protocol

Our ELISA protocol was based on previously published methodologies with modifications [8]. Briefly, white 96-well maxisorp microtitre plates (Nunc 436110) were coated overnight at 4°C with 100 µL of 500 ng/mL Spike RBD (GenScript, Z03483) or Nucleocapsid (GenScript, Z03480) protein in 1X Dulbecco’s phosphate-buffered saline (PBS) pH 7.4 (Sigma D1408). Wells were washed three times with PBS containing 0.2% (v/v) Tween-20 (PBS-T), blocked with 100 µL 3% (w/v) BSA in PBS with 0.1% (v/v) Tween-20 for 1 h at room temperature (RT), then washed once with PBS-T, before addition of 50 µL serum diluted to 1:100 in 1% (w/v) BSA in PBS with 0.1% (v/v) Tween-20. Plate washing was performed by repeated plunging of plates into a bucket filled with PBS-T and flicking of well contents into a sink. After 1 h incubation at RT, wells were washed five times with PBS-T and incubated with 100 µL of horseradish peroxidase (HRP)-conjugated anti-human IgG, IgM or IgA antibodies diluted to the optimal concentration in 1% BSA (w/v) in PBS with 0.1% Tween-20 for 1 h at RT. Wells were washed five times with PBS-T then 100 μL of Super Signal ELISA Pico enhanced chemiluminescent (ECL) substrate (Pierce, Rockford, IL, USA) was added and light emission (stable after 1 min) was measured using a Victor-Nivo luminescence plate reader. For high throughput screening of samples, steps downstream of sample addition were automated as outlined in the supplementary materials and methods.

### Statistical analysis

ELISA data was expressed as the normalized Log_10_ emission at 700nm. ROC analysis and cutoffs were determined using GraphPad Prism 8 software. Estimates of seroprevalence were calculated using R with 95% confidence intervals were calculated by bootstrapping. Bayesian analysis to determine the probability of positivity for each sample was determined using R based on the distributions of the positive and negative values described as mixed distributions. Full details of statistical analysis and R codes are given the Supplementary Materials and Methods.

## Results

### Optimization of manual and automated ELISA protocol conditions

To optimize our assay we used a library of 184 serum samples collected pre-2020 as negative controls, and a panel of 43 sera from individuals infected with SARS-CoV-2 as positive controls. Initial optimization of assay conditions was carried out with defined pools of sera from 5 positive donors and 5 negative donors. Noting that even small gains in specificity can substantially reduce the number of false negatives in large sero-surveys, we optimized the concentration and amount of antigen used for coating, blocking and washing conditions. Overall, we found that the principal factors affecting assay performance were the coating conditions and the necessity of stringent washing (Table S1 and Figure S1).

To handle large numbers of samples we optimized our ELISA assay for automation. We investigated the use of an enhanced chemiluminescence (ECL) substrate as these substrates have superior sensitivity compared to traditional colorimetric absorbance substrates such as *O*-phenylenediamine dihydrochloride (OPD) [9] and do not require a stopping step facilitating automation. Comparing different protocols to distinguish our responses to the N antigen in positive and negative donors we determined that ECL was marginally superior to OPD with a larger separation between positive and negative control values (Figure S2 A and B). Importantly, conducting the analysis on a robotic platform did not compromise assay sensitivity and specificity (Figure S2 C-D).

### Combing IgG responses to multiple antigens gives optimal sensitivity and specificity

Having established optimal ELISA conditions, we wanted to determine the optimal antigen, or combination of antigens for seroprevalence surveys. We therefore compared responses to the full S1 domain of the Spike protein (S1), Nucleocapsid protein (N) and the receptor binding domain (RBD) of the Spike protein. Overall, the sensitivity and specificity of responses to the N and RBD were comparable, with the N protein being slightly superior (Figure 1A and B). Surprisingly, the S1 protein gave very poor sensitivity and specificity with many negative samples giving high values (Figure S3). We next investigated the possibility of combining data for both antigens. Plotting responses to the RBD and N antigens revealed that even the less responsive positive control samples generally had at least elevated responses to both antigens (Figure 1C). Thus, using the mean of the responses to the RBD and N responses, we found that a cutoff of 1.302 gave 100% sensitivity and 98.91% specificity (Figure 1D). Neither IgA nor IgM responses distinguished positive and negative donors as well as IgG, and averaging IgA or IgM responses to both antigens did not substantively improve the assay (Figure S4).

**Figure 1:**
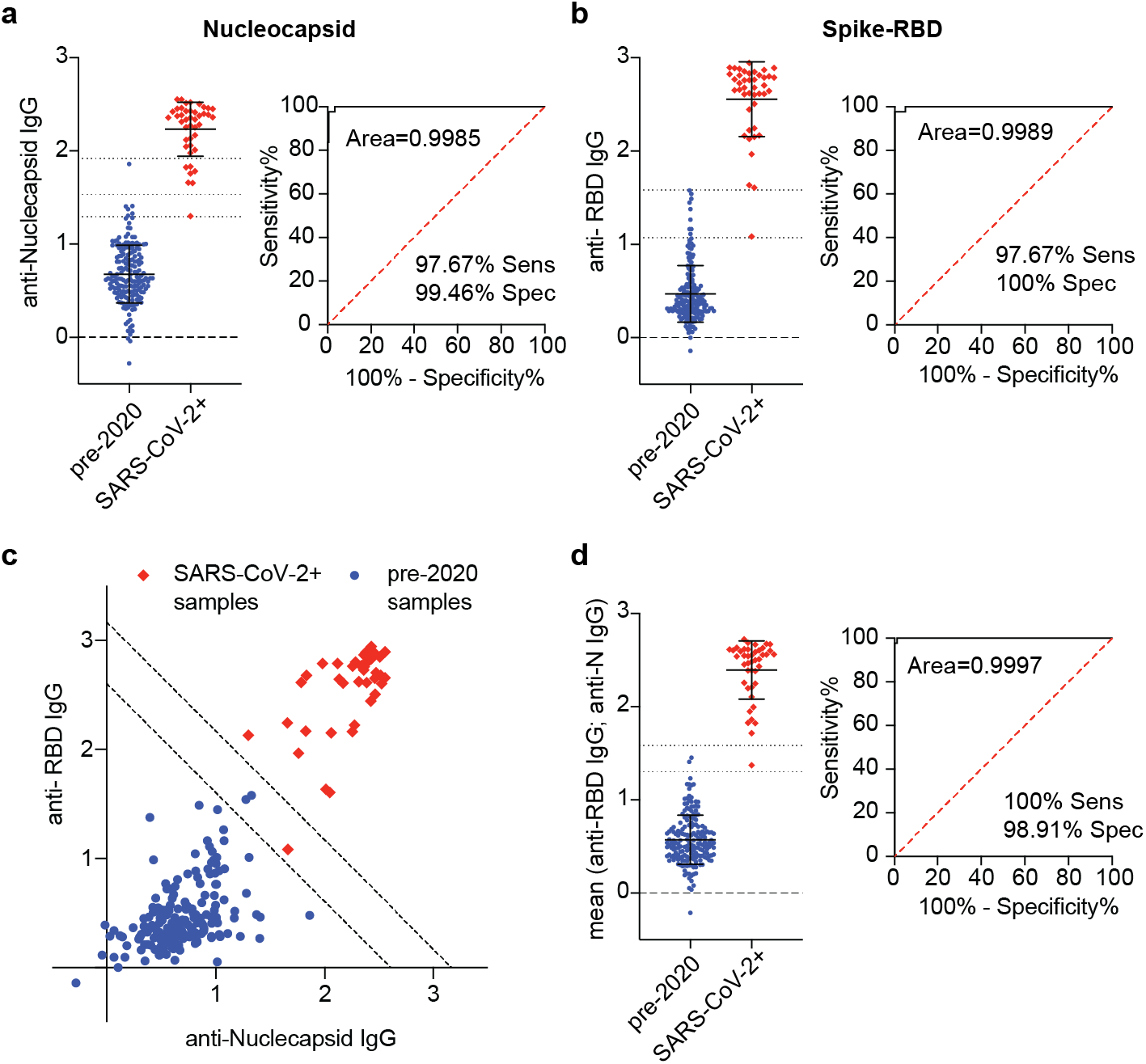
Combining IgG responses to different antigens improves sensitivity and specificity. IgG responses to the N antigen (a) and RBD antigen (b) among positive and negative control samples and corresponding ROC curve used to determine the 100% sensitivity and specificity cutoffs for ELISAs using that antigen (dashed black lines on graph); individual data and mean ± SD shown. (c) Relationship between responses to the N and RBD antigens among positive and negative control samples, dashed lines represent the 100% specificity and sensitivity cutoffs derived from the mean of the IgG responses to the N and RBD antigens. (d) Mean responses to the N and RBD antigens among positive and negative control samples and corresponding ROC curve.

### The seroprevalence of SARS-CoV-2 is low in Australia

We next used our dual-antigen IgG ELISA to assess the seroprevalence of SARS-CoV-2 infection among 2991 individuals, providing blood samples at 10 hospital sites across 4 states in Australia in May and June 2020. These individuals were enrolled in a prospective cohort study to determine the prevalence of asymptomatic SARS-CoV-2 infection in individuals undergoing elective surgery in Australia; full demographic information is given in the full manuscript describing this prospective cohort study separately (Coatsworth *et al*. Submitted). In our initial screen 41/2991 were above our cutoff of 1.302 (Figure 2A), correcting for the specificity of our assay we calculated the seroprevalence to be 0.28% (0 to 0.71%). To confirm our positive results, we retested the top 2.7% of samples from each site in parallel with our complete set of positive and negative control samples. In this analysis 15 individuals remained above the 100% specificity cutoff (Figure 2B), however plotting the RBD and N values showed that only 5 samples were strongly positive for both antigens, clustering with our positive controls. In contrast the remaining 10 putative positives were close to the cutoff and were in many cases strongly positive for only one or other antigen, thus we reasoned these might be false positives. Of note, 1/5 (20%) of our high-confidence positive samples was a contact of a known SARS-CoV-2+ individual, compared to 14/2986 (0.47%) in the remainder of the cohort (p= 0.0248 by two-tailed Fisher’s exact test; odds ratio=53.1 (4.07-357) giving us confidence that our assay was detecting true positive individuals.

**Figure 2.**
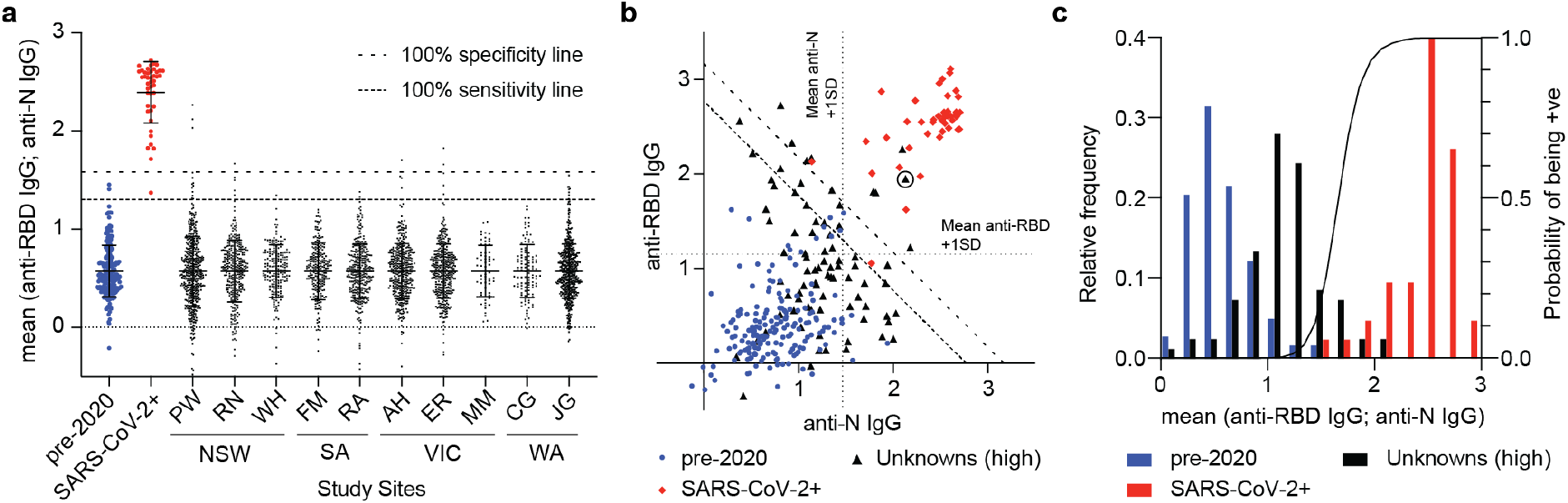
Estimation of seroprevalence of SARS-CoV-2 in Australia. (a) Normalized averaged responses to the RBD and N antigens for each of the 2991 individuals in the study separated by study site and state. (b) Anti-N and anti-RBD responses for the top 2.7% samples from each site (n = 80) compared to the positive and negative controls; the circled unknown sample was a contact of a SARS-CoV-2+ individual. (c) Frequency distribution of the negative, positive and unknown samples (bars) plotted against the calculated probability of positivity in a Bayesian model based on the distributions of the positives and negative samples.

To avoid biases associated with the use of cutoffs we also calculated the probability of each of our 80 retested samples being positive based on the known distributions of the positive and negative results (Figure 2C). This analysis determined that the top 6 samples each had a >50% (58-99%) probability of being positive, while the remaining 9 potentially positive samples had individual probabilities of being positive of 10-47%. By summing the probabilities of positivity among these samples we can estimate that ∼8 (0.27%) individuals in our cohort would be positive which is similar to our original estimate of seroprevalence.

## Discussion

Here we report results from the first large scale seroprevalence survey in Australia. We estimate a seroprevalence of 0.28%, which - given a population estimate for Australia of 25.50 million individuals - equates to 71,400 infections (95% CI: 0 to 181,050). At start of sample collection (2^nd^ June) 7387 cases/102 deaths had been reported in Australia, rising to 11,190 cases/116 deaths by 17^th^ July when sample collection finished suggesting that testing was capturing 10-15% of cases and that there was a low case fatality rate, similar to other jurisdictions with high testing rates [10]. Note that due to the small number of positive samples we have not attempted to stratify our analysis based on the demographic characteristics of our cohort. A key caveat of our study is that the positive controls used for assay validation are skewed to hospitalized individuals and thus we do not know with certainty the performance characteristics of the assays for asymptomatic cases who are known to have lower antibody levels [4, 11]. Moreover, a recent study has suggested that asymptomatic cases may not always seroconvert, though the assays used there had lower sensitivity than we report for our assay [5, 11]. Overall however, these data suggest that the low case number seen in Australia was reflective of low community transmission not inadequate testing. This is supported by the fact that the subsequent outbreak in Melbourne in July/August 2020 emerged from breaches of hotel quarantine of overseas travelers rather than undetected community transmission.

A variety of assays of have been put forward for the assessment of seroprevalence of antibodies to SARS-CoV-2. Lateral flow devices were used in early studies, but these devices have insufficient sensitivity and specificity for use in low prevalence settings [12]. However, more recent studies using ELISA based assays with greater statistical rigor have overcome some of these issues and given reliable estimates of seroprevalence in higher-transmission areas such as the United States [3, 13, 14]. More recently, commercial electrochemiluminescence-based assays have been developed that offer high degrees of sensitivity and specificity as well as standardization [6, 15]. However, these assays only assess IgG responses to a single antigen and are relatively expensive. By combining results from responses to antigens and using convergent statistical approaches we show how an assay that can be established in ordinarily equipped laboratories can obtain credible estimates of SARS-CoV-2 seroprevalence, even in low transmission settings.

## Data Availability

All data pertaining to this paper is reported in the manuscript.

## Acknowledgements

The authors acknowledge the contributions of all participants, study sites, local investigators, surgical and anaesthetic teams, the study coordinators Sophie Wallace and Lucy Morris.

